# *Vibrio Cholerae* O1 Transmission in Bangladesh: Insights from a Nationally-Representative Serosurvey

**DOI:** 10.1101/2020.03.13.20035352

**Authors:** Andrew S Azman, Stephen Lauer, M. Taufiqur Rahman Bhuiyan, Francisco J Luquero, Daniel T Leung, Sonia Hegde, Jason Harris, Kishor Kumar Paul, Fatema Khaton, Jannatul Ferdous, Justin Lessler, Henrik Salje, Firdausi Qadri, Emily S Gurley

## Abstract

**Background:** Pandemic *Vibrio cholerae* from cholera-endemic countries around the Bay of Bengal regularly seed epidemics globally. Without reducing cholera in these countries, including Bangladesh, global cholera control may never be achieved. Little is known about the geographic distribution and magnitude of *V. cholerae* O1 transmission nationally. Here we use recent advances in cholera seroepidemiology to describe infection risk across Bangladesh overcoming many of the limitations of current clinic-based surveillance.

**Methods:** We tested serum from a nationally-representative serosurvey in Bangladesh of 2,778 participants with eight *V. cholerae*-specific assays. Using these data with a previously validated machine learning model we estimate the annual seroincidence rate and use Bayesian geostatistical models to create high-resolution national maps of infection risk.

**Findings:** We estimate a national seroincidence rate of 19.1% (95% CI 12.2-26.9%). Our high-resolution maps reveal large heterogeneity of infection risk, with community-level annual infection risk within the sampled population ranging from 2.4-69.0%. Across the country, we estimate that 31.0 million (95% CI 19.8-43.7 million) occurred in the year before the survey with most of these infections occurring in urban areas, including Dhaka, the capital.

**Interpretation:** Serosurveillance provides an avenue for identifying areas with high *V. cholerae* O1 transmission and exploring key risk factors for infection across geographic scales. This may serve as an important tool for countries to plan and monitor progress towards 2030 cholera elimination goals.

## Introduction

As the 7th cholera pandemic approaches its 60^th^ year, the global public health community is making plans to end cholera as a public health threat by 2030 through reducing transmission and nearly eliminating cholera mortality.^1^ However, these aspirations are challenged by new lineages of pandemic *Vibrio cholerae* O1 that repeatedly arise in South Asia and spread worldwide seeding local and regional epidemics that can last decades, including massive contemporary outbreaks in Haiti and Yemen.^2–5^ Without a significant reduction in cholera transmission in highly endemic countries around the Bay of Bengal, like Bangladesh, *V. cholerae* dissemination to other parts of the world may continue and global cholera control may never be achieved.

Despite years of research on cholera in Bangladesh, little is known about the magnitude and spatial distribution of pandemic *V. cholerae* transmission across the country. Most surveillance efforts have focused on just a few sites within the country, including the sentinel icddr,b hospital in Dhaka and the demographic health and surveillance site in Matlab.^6^ The Ministry of Health & Family Welfare of Bangladesh is developing a national cholera control plan aimed at making significant reductions in cholera morbidity and mortality in the next decade. With more than 164 million people living within the country, targeted interventions focusing on high risk populations are needed. However, most patients with acute watery diarrhea are not systematically tested for cholera and there is no systematic reporting of suspected cholera nationally; leaving decision makers with few data points to devise plans or to monitor progress.

Recent advances in cholera seroepidemiology have shown that cross-sectional antibody profiles can be used to identify individuals infected by *V. cholerae* O1 in the last year, thus providing a measure of the annual infection incidence rate.^7^ Combined with these analytic methods, serosurveys can be used to gauge the extent of recent transmission in a population. These estimates of recent transmission can be used to prioritize resources, track progress in the fight against cholera and to identify high-risk populations for targeted interventions. Data to inform program implementation and monitor progress would make Bangladesh’s efforts to control cholera more efficient and serve as a proof-of-concept for a tool that could be applied globally. Here, we use samples from a nationally representative serosurvey to estimate the annual infection incidence rate in Bangladesh, to identify factors associated with *V. cholerae* O1 infection and to identify areas of the country where transmission risk is the highest.

## Methods

### Study Population and Survey Design

We used data from a previously described two-stage (household and community) cluster survey originally designed to estimate dengue seroprevalence in Bangladesh.^8^ In brief, from December 2015 through January 2016, 70 communities from a total of 97,162 in the 2011 national census were selected with probability proportional to each community’s population. In rural areas, communities were defined as villages, whereas in urban areas they were defined as wards (i.e., neighborhoods). In each selected household, study staff identified the household head, described the study and invited them to participate in the study. If the household head agreed to participate, all household members older than 6 months of age were invited to take part. Within each community, study staff continued enrolling households until at least 40 serum samples were collected from at least 10 households. Within each household, study staff administered both individual and household questionnaires with questions about household-level infrastructure, wealth and assets and individual data on demographics and travel history as well as collecting 5ml venous blood (3ml from children ≤3) from all consenting individuals.

### Laboratory Methods

Aliquots of serum samples were frozen at −80ºC after collection and transport to icddr,b (Dhaka, Bangladesh). All lab analyses were conducted at icddr,b from May through October 2018. Vibriocidal titers were estimated with standard methods using *V. cholerae* O1 Ogawa (X-25049) or Inaba (T-19479) as the target organisms with anti-Lipopolysaccharide (LPS) monoclonal antibodies used for quality control. The complete protocol for the vibriocidal analyses, including rejection criteria and interpretation is available online at https://www.protocols.io/view/bangladesh-national-serosurvey-vibriocidal-protoco-6ydhfs6.

We measured serum isotype-specific (IgM and IgG) antibody responses against *V. cholerae* O1 Ogawa and Inaba LPS and cholera toxin B subunit (CTB) using standardized ELISA methods as previously described.^9^ We read plates kinetically at 450 nm for 5 min and normalized the maximum rate of change in optical density in milliabsorbance units per minute across plates by calculating the ratio of the test sample to a monoclonal antibody standard included on each plate. ELISA samples were rejected and rerun if any of the three sample wells were greater than two times or less than half the median of the three wells.

### Statistical Analyses

Our primary objective is to estimate the proportion of the population with a meaningful immunologic exposure to V. cholerae O1 within the previous year, referred to throughout as the seroincidence rate (or seropositive when describing an individual). To estimate seroincidence, we used a previously published random forest model that identifies recently infected individuals using age, sex, vibriocidal titers (Ogawa and Inaba), anti-LPS IgG and IgA antibodies (maximum of Inaba and Ogawa for each person) and anti-CTB IgG and IgA antibodies.^7^ As a less sensitive and less specific alternative, we used the vibriocidal test to classify individuals following historical convention whereby those with a vibriocidal titer (maximum of Inaba or Ogawa) ≥320 were classified as positive. In supplementary analyses (supplement), we also estimated the 100- and 200-day seroincidence rates.

As the time between sampling an individual and their last infection increases, the sensitivity of the random forest and vibriocidal tests tend to decrease. We model this time-varying sensitivity using a logistic regression with a cubic polynomial for time since an individual’s infection (up to 1 year post-infection) with data from a previously published cohort study.^7,10^ We then we combined the specificity of each test and the time-varying sensitivity, assuming constant risk in the previous year, to adjust our estimates of seroincidence in sampled communities. To estimate seroincidence in the sampled population while correcting for test sensitivity and taking into account the sampling design, we applied a Bayesian hierarchical model, implemented in Stan.^11^ Full details of the seroincidence models are provided in the supplemental appendix.

To extend estimates from the sampled population to the full population of Bangladesh we fit a logistic regression model with distance to the nearest major water body and population density^13^ as covariates and a spatial random field assuming a Matern spatial covariance model. We fit this model to grid cell-level seroincidence estimates from the Bayesian hierarchical model described above. We estimated the joint posterior density of seroincidence at a 5km by 5km grid-cell level across Bangladesh using integrated nested Laplace approximations (INLA) as implemented in the R-INLA package.^12^ We divided the grid cell-level estimates of seroincidence across the country by the population weighted mean to obtain a map of relative infection risk. To estimate the number of infections in the year preceding the survey, we multiplied estimates of the seroincidence rate by the estimated 2015 population^13^ in each grid cell. We report the posterior median and 95% credible intervals from these models (supplemental appendix).

We used leave-one-out cross validation to check the out-of-sample performance of the map predictions. Each grid cell with a sampled community was alternatingly held out, while the INLA model was fit on the remaining grid cells. The INLA model was then used to predict the held-out grid cell. These predictions were compared to a naïve model that predicted the mean of the other sampled grid cells using mean absolute error.

We explored the associations between seropositivity and a number of individual-, household- and community-level risk factors using spatial logistic regression models with random effects for households and communities. All individual and household-level data were collected directly from the survey questionnaires. Population density,^13^ travel time (to the nearest city),^14^ distance to a major water body, altitude and a poverty index^15^ were from publicly available sources. We included spatial correlation using a Matern covariance model and estimated the joint posterior density using INLA.^12^ Odds ratios and corresponding 95% highest posterior density intervals (referred to as 95% CI throughout) were based on 1,000 draws from the posterior. In sensitivity analyses we included models that had either household-level or community-level random effects with or without the Matern spatial covariance function.

All analyses were performed in R. Data and source code to reproduce analyses are available at https://github.com/HopkinsIDD/Bangladesh-Cholera-Serosurvey with additional details provided in the supplemental appendix.

### Ethics and role of the funding source

This study was approved by the icddr,b ethical review board (protocol number PR-14058) and this secondary analysis was reviewed and deemed exempt from review by the Johns Hopkins Bloomberg School of Public Health Institutional Review Board. All adult participants provided written informed consent to participate in the study. Parents/guardians of all child participants provided written, informed consent on their behalf.

The funder of the study had no role in study design, data collection, data analysis, data interpretation, or writing of the report. The corresponding author had full access to all the data in the study and had final responsibility for the decision to submit for publication.

## Results

We collected and tested serum samples from 2,930 individuals coming from 707 households in 70 communities throughout Bangladesh. Study participants were generally reflective of the age-sex distribution of the population, although young children were underrepresented (Figure S1). Overall, 19.5% of the sampled population had vibriocidal titers ≥320, a common yet imprecise threshold to classify individuals as having been infected with V. cholerae O1 within the last year.7,16 Applying the machine learning algorithm we estimate that 20.4% (95% CI 15.1-26.4%) of the sampled population was infected in the year before the survey (referred to throughout as seroincidence).7 Through extending these estimates to the full Bangladesh population, accounting for spatial correlation in risk, we estimate a national seroincidence rate of 19.1% (95% CI 12.2-26.9%), with alternative models producing similar results (supplementary appendix).

Within households, seroincidence ranged from 0-100%, with 339/707 (47.9%) of households having no seropositive individuals. Members of households with at least one other seropositive individual had a 1·7-fold (mean: 1·70, 95% CI: 1·46-1·98) increase in the risk of being seropositive compared to those with no other seropositive individuals in the household. Seroincidence varied across sampled communities with 9 (of 70) communities having a seroincidence rate of less than 5% and 5 communities with more than 50% (Figure 1).

**Figure 1:**
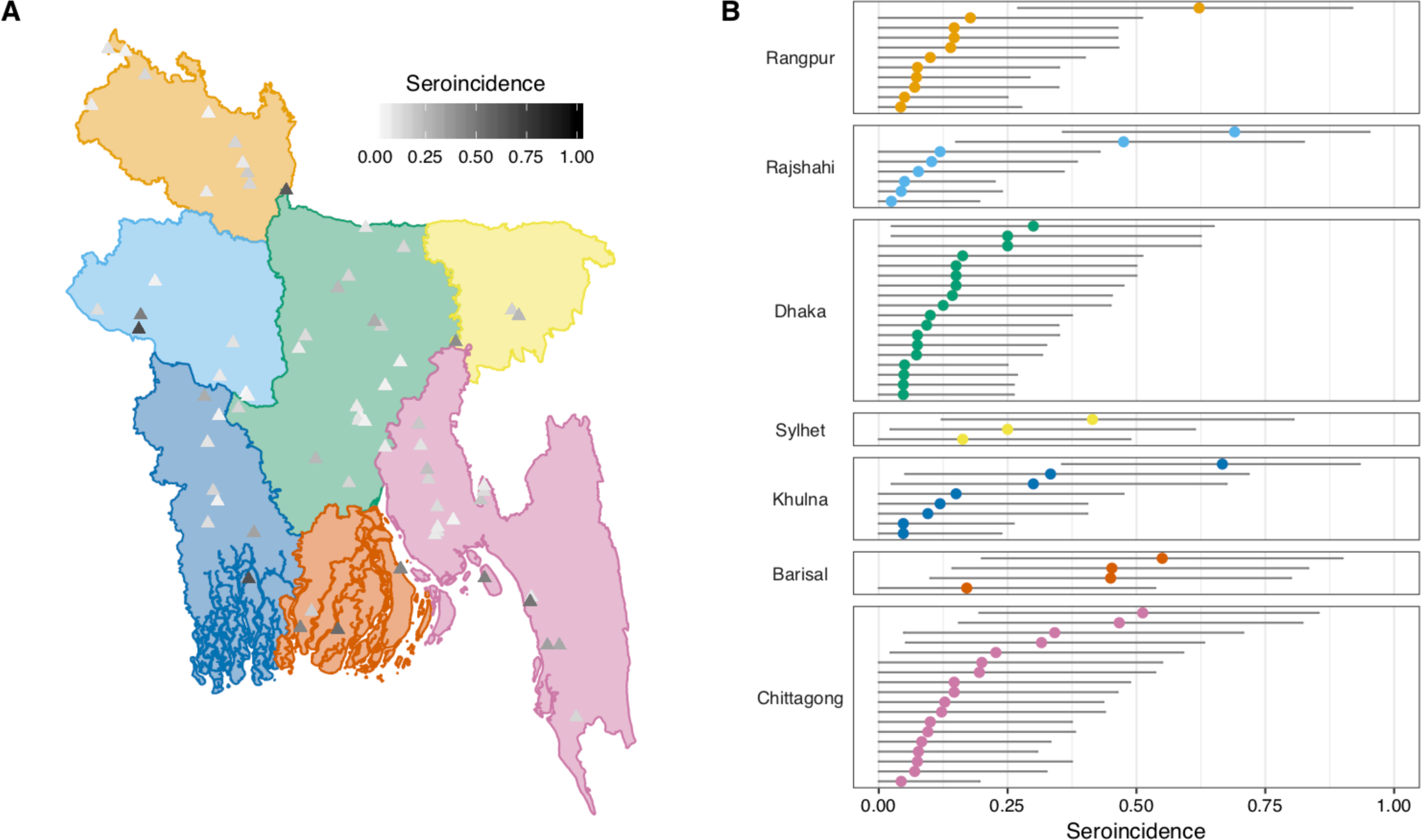
Location and seroincidence of sampled communities by division. Panel A illustrates the location of each community with colors representing each of the seven divisions of Bangladesh and triangle shading representing the seroincidence. Panel B illustrates presents the seroincidence estimates and accompanying 95% confidence intervals for each community, grouped by division (color).

Using a random effects logistic regression model with spatially correlated errors, we estimated the association between a number of individual-, household- and population-level variables and recent (unadjusted) cholera infection (Table 1). Individuals less than five years old had 4.2 times lower odds (aOR 0.24, 95% CI 0.09-0.56) of seropositivity than those 15 years and above, with no differences between males and females (aOR 1.07, 95%CI 0.89-1.28). We found no evidence of an association between recent travel history and the odds of seropositivity. While population density was not significantly associated with seropositivity, in adjusted analyses, we found that individuals living in areas classified as urban by the 2011 census had a 36% (aOR 1.36, 95% CI 1.01-1.84). Alternative models considered, including univariate models and those without spatial correlation, led to similar estimates of the relationship between cholera risk and potential risk factors (Figure S6).

**Table 1.**
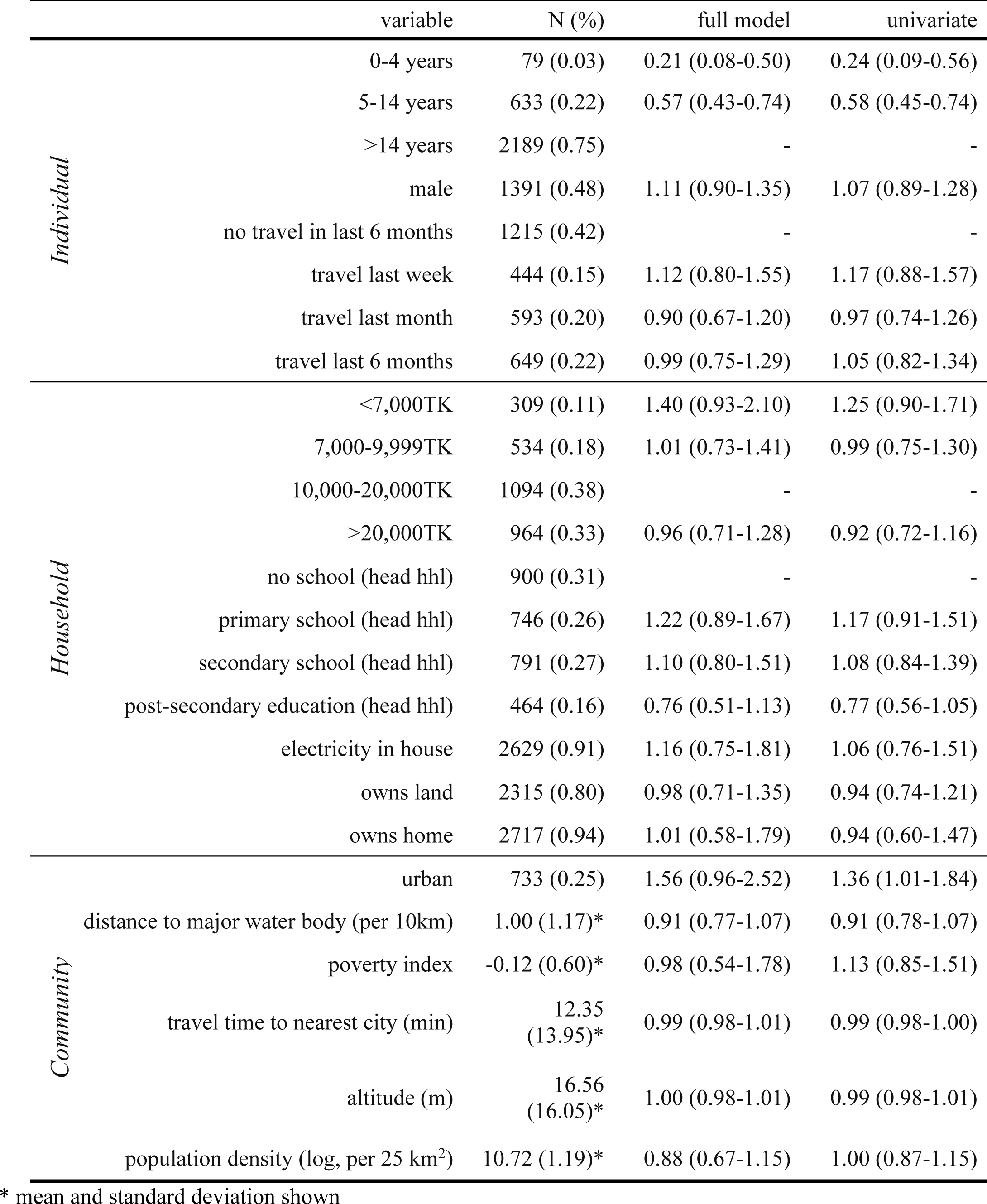
Estimated associations (odds ratios and 95% confidence intervals) between individual-, household- and community-level factors and seropositivity from spatial logistic regression models. The full model includes all covariates shown in table, random effects for household and community in addition to a Matern spatial correlation function. Reference categories for categorical variables are indicated with a hyphen.

In leave-one-out cross validation tests of our national maps of seroincidence, we found that our predictions had little-to-no bias (bias = −0.015), a mean absolute error of 0.093 and 18% less error than a naïve model. Our maps reveal high levels of *V. cholerae* O1 infection risk heterogeneity, with community-level (i.e., grid-cell) relative infection risks ranging from 0.16 to 4.02 (Figure 2). We find that the highest risk of infection in Bangladesh occurs around the Bay of Bengal, including the peninsula, with pockets of high relative risk estimated in the central-west, central-east and north. We estimate that 31 million *V. cholerae O1* infections (median 31.0 million; 95% CI 19.8-43.7 million) occurred in the year before the survey with most of these infections occurring in urban areas, including Dhaka, Khulna, Chittagong and Rajshahi (Figure 2B).

**Figure 2:**
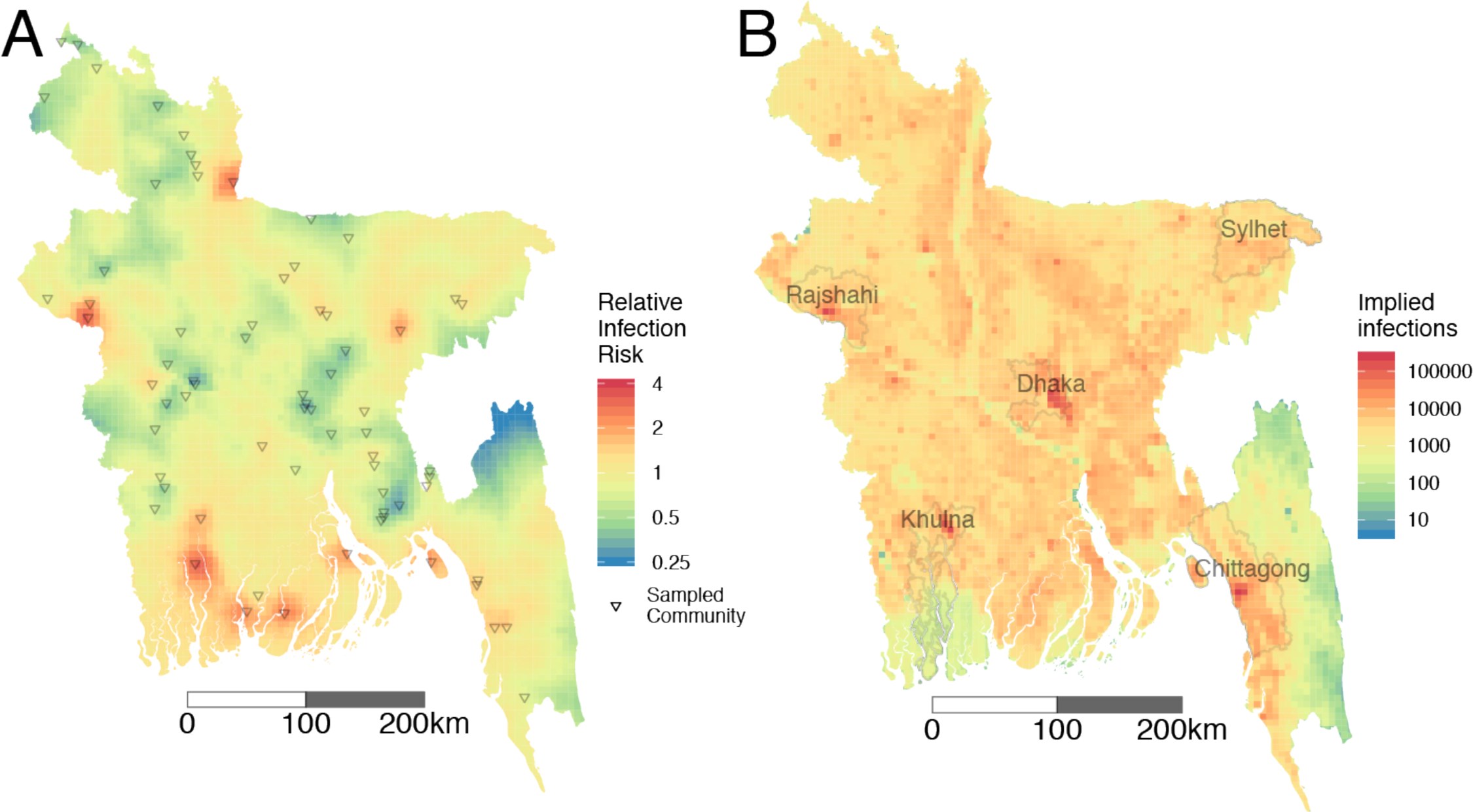
Relative risk of *V cholerae* O1 infection estimated at a 5km x 5km grid cell level (A) and estimates of the number of infections per 5km x 5km grid cell across Bangladesh. **(B)**. Sampled sites shown in triangles in A and five most populous cities in Bangladesh shown with highlighted polygons and labels in B. Estimates are based on using random forest model with population density and distance from water as covariates with the proportion positive at each grid-cell adjusted for the performance of the test.

## Discussion

We found evidence that 1 in 5 people across Bangladesh were infected with *V. cholerae* O1 in the year before the survey with variability in risk between households and communities. Our serology-based approach to estimating the magnitude and geographic variability in cholera risk illustrates a new avenue for understanding cholera epidemiology without the traditional biases related to care seeking behavior and imperfect reporting.

Our results highlight the differences between relative risk and absolute infection burden in Bangladesh. Dhaka is often considered as one of the highest risk areas for cholera in Bangladesh, perhaps due to the presence of a famous reference cholera hospital (the icddr,b hospital) and numerous cholera-related research studies.^18^ While there is likely large variation in infection risk within Dhaka, we found that relative to other areas, the annual risk of infection in Dhaka was 50.3% lower than the average national risk (9.6 vs. 19.1% seroincidence).

However, given the large population of Dhaka, this still translates to a massive infection burden for the city with more than 1.5 million annual infections. Prioritizing cholera control interventions in Dhaka might be well justified despite it not having the highest relative risk due to Dhaka’s role as a major transport and migration hub and the large number of infectious people within the city.

While our results highlight the utility of serosurveys for understanding the overall magnitude of transmission and geographic distribution of risk, there are a number of limitations related to their generalizability and public health utility. As with any diagnostic test, the precision of our seroincidence estimates is limited by the sensitivity and specificity of the random forest model (supplementary appendix). Our random forest model was trained on data from a cohort of severe cholera cases and uninfected contacts in Bangladesh, with few young children and no individuals over 60 years old. Vibriocidal antibody kinetics appear to decay faster than adults,^7,19^ which could lead to underestimation of seroincidence among young children in our study. Future studies describing the post-infection kinetics of mild and asymptomatic *V. cholerae* infections in addition to infections in young children and the elderly will help improve the generalizability of this recent infection model.

Our estimates of seroincidence are high but consistent with previous longitudinal household studies in Bangladesh.^20–22^ For example, one study in Dhaka followed confirmed cholera cases serologically after infection and found that 30% of them had evidence of re-infection in the subsequent year.^20^ Published estimates of the proportion of infections that result in clinical disease range widely, with some suggesting over 100 *V. cholerae* infections for each severe cholera case and others suggesting 4 infections per severe case.^16,23,24^ This ratio likely varies by infecting strain, setting and the ingested inoculum. Future studies gathering clinical surveillance, health seeking behavior and serology data across areas with different epidemiological/transmission characteristics can help us better estimate the true distribution of this ratio and help translate infection incidence to public health burden.

These nationally representative data provided an opportunity to gain new insights to guide the design of future serosurveys related to cholera. While the raw antibody results primarily drive our estimates of seroincidence, corrections for test performance based on time since infection can have a significant effect. Depending on the primary purpose of a serosurvey, models and sampling design can be tailored to maximize test performance. For example, if the aim is to estimate the true size of an outbreak, conducting a serosurvey just after the outbreak, or serial serosurveys during protracted outbreaks may lead to increased sensitivity and more precise estimates of risk. Due to its moderate specificity, this approach is unlikely to be able to verify the absence of cholera transmission in an area. Even if tuned to maximize specificity, the lower limits of detection would be well above one infection per 1,000 population, which is too high in most settings to declare a community cholera-free. The addition of new markers that can help discriminate between proximal and distal infections may improve model performance ultimately bringing the specificity closer to 100% while maintaining adequate sensitivity.

In summary, this nationally representative serosurvey highlights the high proportion of Bangladesh’s population infected annually with *V. cholerae* O1 and identifies potential target areas - both populations with higher risk for infection and higher numbers of total infections - for efficient deployment of control measures, including vaccines. Serosurveillance can be an important tool for tracking changes in transmission, and impact of interventions, over the coming years in Bangladesh and globally. Efforts to improve the ease of deploying these studies and testing of samples would further increase their value.

## Data Availability

Data and source code to reproduce analyses are available at https://github.com/HopkinsIDD/Bangladesh-Cholera-Serosurvey

## Author Contributions

ESG, HS and KKP conducted the original survey. ESG, HS and ASA conceptualized this study. FQ, MTRB, FK, JF and FQ were responsible for the laboratory testing. DTL and JH assisted with reagents, laboratory QA/QC and the documentation of the vibriocidal protocol. ASA was responsible for the funding for this study. ASA, SL, FJL and JL conceptualized the analyses. ASA and SL analysed the data. ASA and SL wrote the first draft of the manuscript. All authors participated in writing and reviewing the manuscript.

